# Functional components of a community of practice to improve community health worker performance: A qualitative study

**DOI:** 10.1101/2021.09.21.21263835

**Authors:** Rachel Hennein, Joseph M. Ggita, Patricia Turimumahoro, Emmanuel Ochom, Amanda J. Gupta, Achilles Katamba, Mari Armstrong-Hough, J. Lucian Davis

## Abstract

**Background:** Communities of Practice (CoPs) are a low-cost strategy for health workers to share and create knowledge through social interactions to improve the delivery of high-quality care. However, there remain critical gaps in understanding the behavioral mechanisms through which CoPs can facilitate community health worker’s (CHWs) performance. Therefore, we carried out a qualitative study of a CoP to identify the behavior change techniques (BCTs) and intervention functions that improved CHWs’ performance.

**Methods:** We organized CHWs from two tuberculosis (TB) clinics in Kampala, Uganda into a CoP from February to August 2018. We conducted interviews with CoP members to understand their perceptions of how the CoP influenced delivery of TB contact investigation. Using an abductive approach, we first applied inductive codes characterizing CHWs’ perceptions of how the CoP activities affected their delivery of contact investigation. We then systematically mapped these codes into their functional categories using the BCT Taxonomy and Behavior Change Wheel framework. We triangulated all interview findings with detailed field notes.

**Results:** All eight members of the CoP agreed to participate in the interviews. CHWs identified five CoP activities as improving the quality of their work: (1) individual review of feedback reports, (2) collaborative improvement meetings, (3) real-time communications among members, (4) didactic education sessions, and (5) clinic-wide staff meetings. These activities represented nine different BCTs and five distinct intervention functions. Taken together, CoP meetings enabled members to foster social support, problem solving, and knowledge sharing. The CoP enabled CHWs to identify barriers they face in the field and develop solutions. The CoP was motivating, strengthened their social and professional identities within and outside of the group, and improved their self-efficacy.

**Conclusions:** We identified several behavioral mechanisms through which CoPs may improve CHW performance. Future studies should evaluate the importance of these mechanisms in mediating the effects of CoPs on program effectiveness.

## Introduction

Sub-optimal healthcare worker performance is a major barrier to delivery of high-quality health services in low- and middle-income countries (1-3). This barrier is particularly salient for community health workers (CHWs), who have limited formal health professional education and access to training in low-income countries (4). Many quality improvement initiatives include training to improve healthcare worker performance; however, a systematic review of such strategies found that while training was associated with only moderate improvements in performance, when combined with group problem solving, large improvements were observed (3). Furthermore, a Cochrane qualitative evidence synthesis concluded that providing continuous education and enabling CHWs to share their experiences with peers facilitated their work (5).

Communities of Practice (CoPs) offer a promising, yet understudied, mode of delivery for continuous group learning and problem solving (6). Communities of Practice (CoPs) are groups of people with a common work objective who meet regularly to support each other, share and create knowledge, and explore innovations (7). In their original studies among West African tailors, Lave and Wenger (1991) developed the term CoP to describe the organic learning that occurs among tradespersons and other professionals-in-training (6, 8). Over time, health services researchers have explored using CoPs as an instrument of change to improve healthcare worker performance (8-11). However, systematic reviews have identified critical gaps in our understanding of CoPs related to their functional components, mechanisms of action, and fit of CoPs to improve delivery of health services in different settings (7, 9, 10).

Specifying the functional components of CoPs will enable practitioners to design better strategies to devise and establish them for continuous quality improvement (12, 13). Functional components are aspects of the intervention that elicit behavior change. Because these concepts are still evolving (11), applying behavioral theory to data collected in empirical studies of CoPs could improve our understanding of when, where, how, and under what conditions these groups can be engineered to improve performance (10, 14). For example, the Behavior Change Technique (BCT) Taxonomy and the Behavior Change Wheel provide comprehensive approaches to cataloguing the functional components of complex health interventions in order to design and implement strategies that optimize outcomes (15, 16).

Therefore, we performed a qualitative study to identify the functional components of a CHW CoP formed to improve tuberculosis (TB) contact investigation in Kampala, Uganda. Through semi-structured interviews and field notes, we aimed to explore CHWs’ experiences participating in the CoP and determine the extent to which the CoP was acceptable, feasible, and effective in facilitating their delivery of contact investigation. We analyzed qualitative data using the Behavior Change Technique (BCT) Taxonomy (16) and the Behavior Change Wheel’s intervention functions (15). In so doing, we aimed to identify behavioral mechanisms to describe how CoP activities function to improve CHW performance in low-resource settings.

## Methods

### Setting

Uganda has a high TB burden, with an annual incidence rate of 201 cases per 100,000 and an annual mortality rate of 26 deaths per 100,000 (17). In Kampala, TB services are provided free of charge through the Uganda National TB and Leprosy Program (NTLP) and Kampala Capital City Authority. CHWs support regular health workers in delivering TB services, with funding and technical assistance from non-governmental or research organizations partnering with the NTLP. CHWs are responsible for community-based treatment adherence support and contact investigation, as well as clinic-based TB symptom screening, education, and counseling. Clinical data is recorded in paper logbooks, or in electronic case-record forms on mobile tablets.

### CoP Intervention

Our research team established a CoP in February 2018 within TB units at two public health centers in Kampala to support CHWs delivering contact investigation. To establish the CoP, the research team encouraged members to meet weekly on Friday mornings to share stories about their experiences, successes, and challenges in delivering contact investigation in the prior week. Leadership responsibilities rotated weekly among all participants, with the designated chairperson assigned at the end of each meeting to organize and lead the discussion the following week. To catalyze discussions, the research team provided performance reports listing incomplete contact investigation records for each CHW. The reports also presented facility-level process indicators for each step of the TB contact investigation cascade, including the stepwise proportions of (1) cases interviewed, (2) contacts screened, (3) eligible contacts completing evaluation, and (4) individuals diagnosed with active TB (18).

### Data Collection

A Ugandan social scientist (JG) prospectively collected field notes during weekly CoP meetings. Field notes summarized meeting content, participation of members, interactions between participants, and meeting tone. Research staff invited all CoP participants to interview at a place and in a language (English or Luganda) of their choice five months after CoP initiation. Two of the researchers (MAH, JG) developed an interview guide to probe how CHWs perceived their roles delivering contact investigation during CoP implementation (Additional File 1). Three Ugandan members of the research team (JG, EO, PKT) who participated in CoP implementation discussed and revised the guide after reviewing the field notes. A Ugandan social scientist (JG) obtained verbal consent and conducted and audio-recorded all interviews. We then de-identified and transcribed all interviews, and translated Luganda interviews into English. The interviewer (JG) revised all transcripts for accuracy. The Makerere University School of Public Health Higher Degrees Research Ethics Committee and the Yale University Human Investigation Committee approved the study. We reported all findings using the Consolidated Criteria for Reporting Qualitative Research (COREQ) checklist (19).

### Data Analysis

A non-Ugandan researcher (RH) with field experience in Uganda coded transcripts in ATLAS.ti using an abductive approach (20). Abductive analysis employs both inductive codes that emerge from the data and deductive codes informed by theory. We followed a three-step abductive analytic process to identify BCTs and intervention functions (Figure 1) (21). This included: (1) cataloging activities, (2) identifying how activities affected behavior, (3) classifying BCTs, and (4) mapping intervention functions. First, the coder analyzed interviews inductively to identify CoP activities that CHWs described as benefitting their work. We cross-referenced activities that emerged from interviews with field notes and specified their actors, participants, modes of delivery, and frequencies using the Template for Intervention Description and Replication (TIDieR) checklist (22). Second, the coder applied inductive codes to describe CHWs’ perceptions on how and why these activities influenced their delivery of TB contact investigation. In the third step, we systematically mapped inductive codes and themes to the BCT Taxonomy. The BCT Taxonomy was created to characterize active ingredients of complex interventions (16, 23). We adapted definitions from the BCT Taxonomy to describe the CoP. We then organized the BCTs by relevant CoP activity and noted contextual factors that facilitated CHWs’ performance. Finally, we mapped BCTs to intervention functions using the Behavior Change Wheel to understand underlying mechanisms through which CoPs influence practice (24). Three authors (RH, JLD, MAH) reviewed and discussed the mapped BCTs and intervention functions to reach consensus. Ugandan team members (JG, EO, PKT) and non-Ugandan researchers with extensive local research experience (JLD, MAH, AJM) reviewed and validated the code structure. We triangulated data from both the field notes and semi-structured interviews. We defined data saturation as the point at which novel inductive codes ceased to emerge from the data (25).

**Figure 1.**
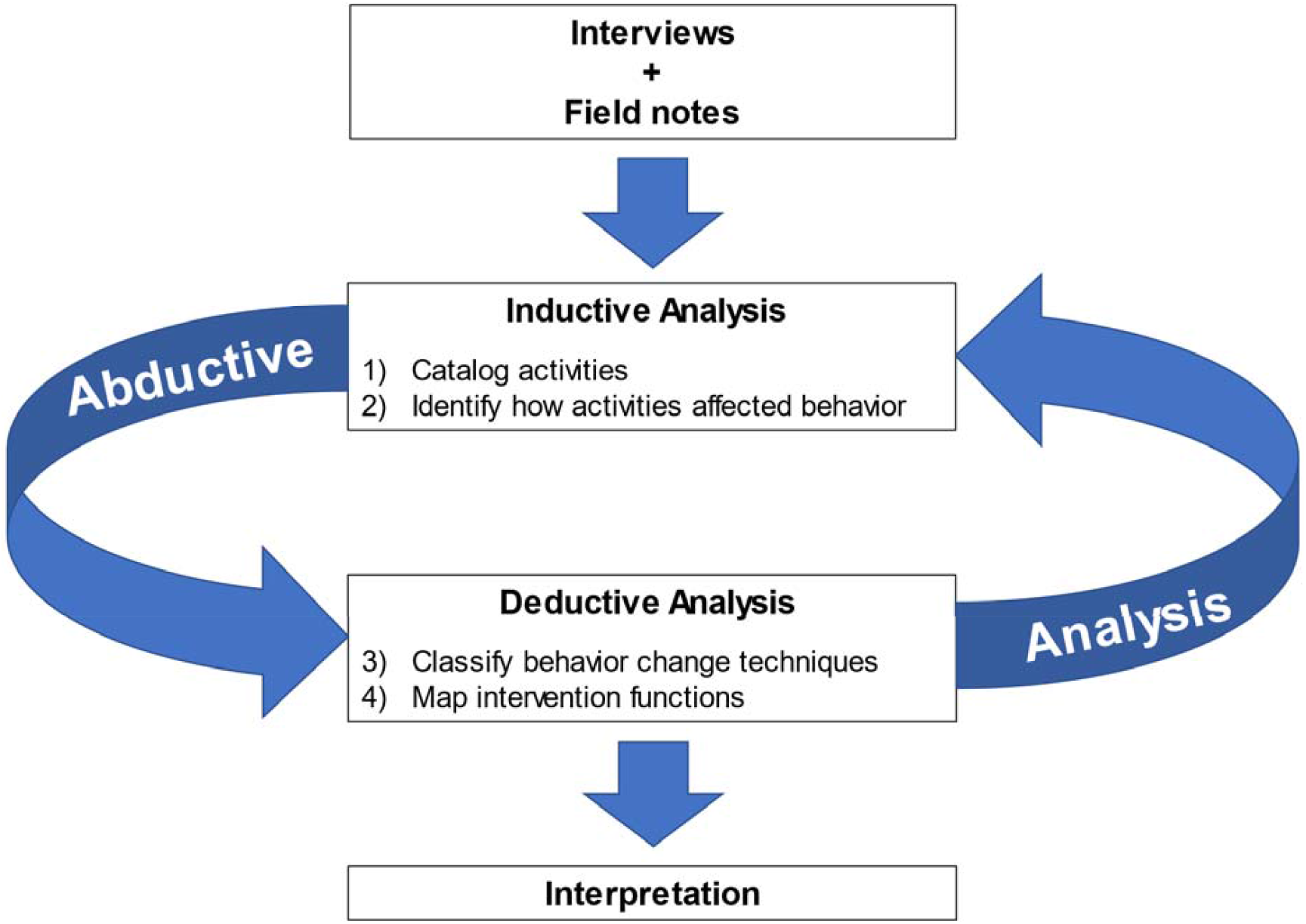
Process model for identifying behavior change techniques and intervention functions of the Community of Practice using abductive analysis.

## Results

### Sample

All eight CHWs from the CoP agreed to participate in interviews. Four interviews were conducted in English, two in a mixture of English and Luganda, and two in Luganda. Interviews lasted from 48 to 69 minutes. Median age of participants was 39.5 years (range 26-51) and six (75%) participants were female (Table 1). We reached data saturation after six interviews.

**Table 1.** Participant characteristics.

| **Characteristics** | **n (%)** |
| --- | --- |
| Gender |  |
| Female | 6 (75) |
| Male | 2 (25) |
| Age range (years) |  |
| 21-30 | 2 (25) |
| 31-40 | 2 (25) |
| 41-50 | 3 (38) |
| 51-60 | 1 (12) |
| Education |  |
| Vocational | 1 (12) |
| O-Level | 1 (12) |
| A-level | 3 (38) |
| Diploma | 1 (12) |
| University | 2 (25) |
| Interview language |  |
| English | 4 (50) |
| Luganda | 2 (25) |
| English and Luganda | 2 (25) |

Abbreviations: A-level, Advanced (secondary school) level; O-level, Ordinary (secondary school) level

### CoP content

CHWs identified five core components from the CoP that facilitated their delivery of household contact investigation services (Table 2). These included: (1) individual review of feedback reports, (2) collaborative improvement meetings, (3) real-time communication among members, (4) didactic education sessions, and (5) clinic-wide staff meetings. We identified relevant BCTs (Table 3) and Behavior Change Wheel functions for each activity discussed (Table 4 and Figure 2). Additional File 2 describes peripheral components that facilitated the delivery of these core components, such as cellphone data plans to participate in real-time communication among members.

**Table 2.** Description of Community of Practice activities that emerged from the interviews.

| **Description** | **Mode of delivery** | **Organized by** | **Participants** | **Frequency** |
| --- | --- | --- | --- | --- |
| *Activity 1: Individual review of feedback reports* |  |  |  |  |
| Reading and reflecting on performance reports that included key indicators for the tuberculosis contact investigation cascade | Hard copy/paper; electronic copy | Research coordinator | Community of Practice members | Weekly |
| *Activity 2: Collaborative improvement meetings* |  |  |  |  |
| Weekly meetings led by a rotating community health worker chairperson for the group to share experiences, review feedback reports with each other, and solve problems together | In-person | Community of Practice champion (Champion rotated weekly) | Community of Practice members | Weekly |
| *Activity 3: Real-time communications among members* |  |  |  |  |
| Text messages and phone calls to request immediate advice or assistance or share concerns | WhatsApp, phone calls | Community of Practice members | Community of Practice members | Continual |
| *Activity 4: Didactic education sessions* |  |  |  |  |
| Lectures and interactive teaching sessions on topics relevant to contact investigation that were organized by the research team and delivered by invited clinicians | In-person | Community of Practice members with help from research staff and clinicians | Community of Practice members | As needed |
| *Activity 5: Clinic-wide staff meetings* |  |  |  |  |
| Meetings with other clinic staff to provide updates on the contact investigation activities being carried out by community health workers | In-person | Community of Practice members | Clinic staff, Community of Practice members | Monthly |

**Table 3.** Adapted definitions for behavior change techniques relevant to Community of Practice activities.

| **BCT Category** | **BCT Label** | **Adapted BCT Definition** |
| --- | --- | --- |
| Feedback & monitoring | *Self-monitoring of behavior* | Establish a method for Community of Practice members to regularly examine and record their own behavior(s) during household visits for contact investigation. |
|  | *Feedback on behavior* | Monitor and provide informative or evaluative comments to the actor on performance of contact investigation (e.g., How frequently was sputum successfully collected if indicated?). |
| Goals and planning | *Discrepancy between current behavior and goal* | Draw attention to differences between the Community of Practice members' household visit metrics (i.e. contact investigation process metrics, aggregated by clinic affiliation) and the goal of completing contact investigation for all clients. |
|  | *Problem solving* | Prompt Community of Practice members to analyze factors influencing the desired outcome of completing household contact investigation and generate strategies for overcoming barriers and/or increasing facilitators. |
| Social Support | *Social support (practical)* | Provide practical help from peers and/or supervisors to improve the performance of household contact investigation. |
| Comparison of behavior | *Social comparison* | Draw attention to the performance of other Community of Practice members in carrying out contact investigation to emphasize similarities to and differences from each individual member's own performance. |
| Antecedents | *Restructuring the social environment* | Change the interactions among the Community of Practice members, supervisors, and/or clinic staff to facilitate household contact investigation. |
| Shaping knowledge | *Instruction on how to perform a behavior* | Advise or agree on how to carry out household contact investigation (includes skills training). |
| Identity | *Identity associated with changed behavior* | Construct a new self-perception as a more skilled community health worker conducting household contact investigation. |

Abbreviations: BCT, behavior change techniques.

**Table 4.** Behavior change techniques for Community of Practice intervention activities linked to Behavior Change Wheel intervention functions.

| Intervention Component | Intervention Content |  |
| --- | --- | --- |
|  | BCT Label | Functions |
| *Activity 1: Individual review of feedback reports* |  |  |
| Program provided feedback reports on performance of household contact investigation (e.g., how frequently were sputum samples successfully collected if indicated) to Community of Practice members. | Feedback on behavior | Enablement |
| Community of Practice members reviewed their data to understand their performance in carrying out contact investigation. | Self-monitoring of behavior | Enablement |
| Community of Practice members viewed discrepancies between their household visit metrics and their goal of completing household contact investigation in full. | Discrepancy between current behavior and goal | Enablement |
| *Activity 2: Collaborative improvement meetings* |  |  |
| Community of Practice met weekly to discuss challenges and devise solutions. | Problem solving | Enablement |
| Community of Practice members offered practical support to each other based on challenges discussed. | Social support (practical) | Enablement |
| Community of Practice members compared and discussed their own performance to that of their peers. | Social comparison | Modelling |
| Research staff gave Community of Practice members more decision-making power during the weekly meetings. | Restructuring the social environment | Environmental restructuring |
| *Activity 3: Real-time communications among members* |  |  |
| Program staff created a WhatsApp group for support | Social support (practical) | Enablement |
| *Activity 4: Didactic education sessions* |  |  |
| Community of Practice members identified gaps in their own knowledge and skills and requested appropriate education and training sessions. | Instruction on how to perform a behavior | Training, Education |
| Community of Practice members gained self-efficacy and confidence to engage with communities as health workers. | Identity associated with changed behavior | Training, Education |
| *Activity 5: Clinic-wide staff meetings* |  |  |
| Community of Practice members were recognized for their contributions in the clinic. | Restructuring the social environment | Environmental restructuring |
| Community of Practice members received practical support from clinic staff. | Social support (practical) | Enablement |
| Community of Practice members shared problems with clinic staff to devise solutions. | Problem solving | Enablement |

Abbreviations: BCT, behavior change techniques.

**Figure 2.**
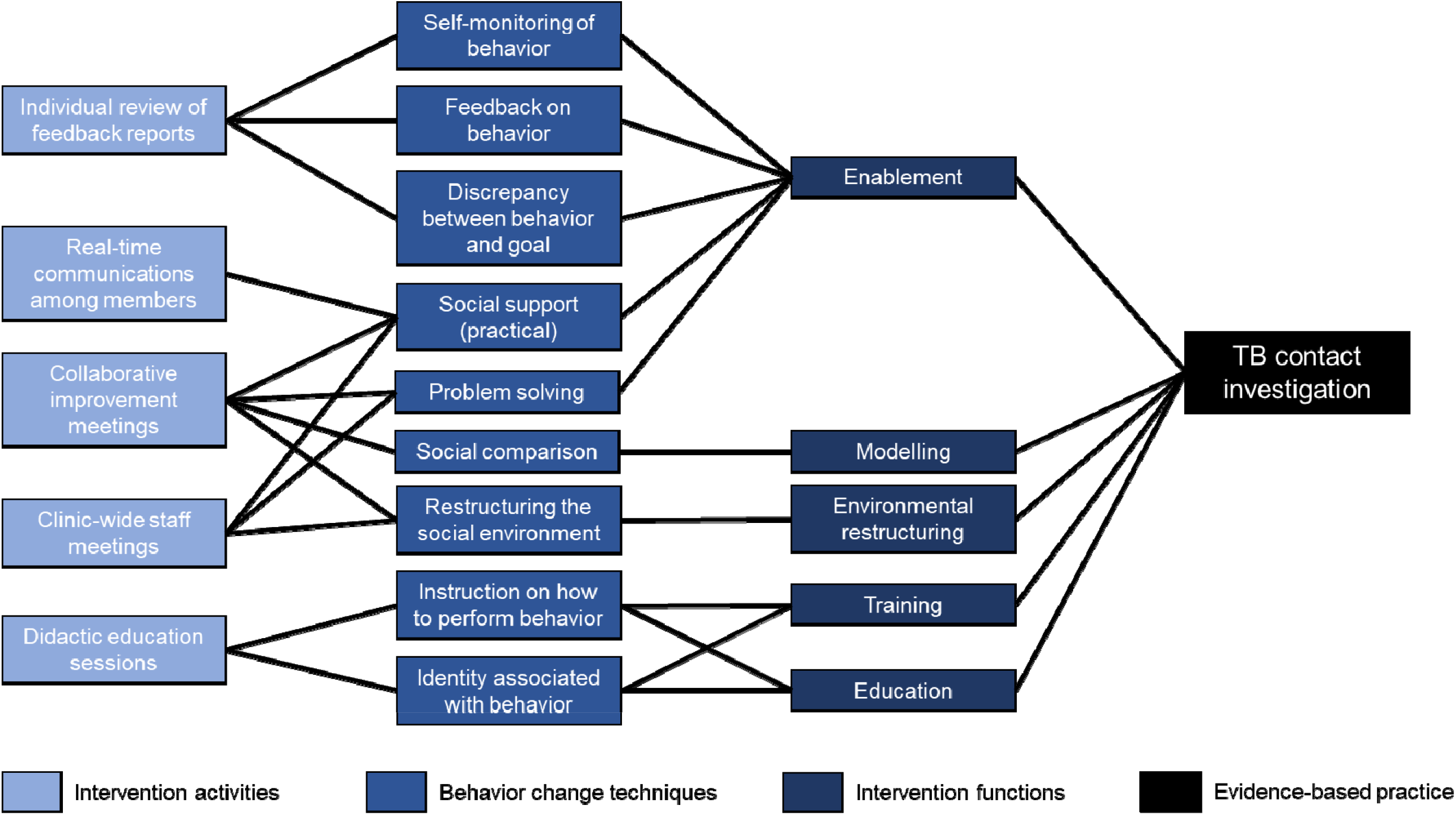
Conceptual model illustrating our implementation mapping exercise for the Community of Practice intervention. Starting at the far left, we linked intervention activities to specific behavior change techniques and related intervention functions, all to facilitate implementation of the evidence-based practice of tuberculosis contact investigation.

### Activity 1: Individual review of feedback reports

First, CHWs stated that review of feedback reports helped them improve the quality of contact investigation services. The research staff provided weekly performance reports to CoP members that included key performance indicators for the TB contact investigation cascade for each site. The weekly reports also included an itemized list of missing case record forms and the associated contacts for each individual CHW. CHWs could review their own individual reports to monitor their performance over time. One CHW explained:

> *“Those reports were very helpful in a way that it helped me figure out my weaknesses, where I had not done well. It would help me know the home visits I have and those I missed so I would know that I am demanded [responsible for] three home visits which was very helpful*.*”*

Many respondents also suggested that reviewing and later referencing feedback reports served as “reminders” to complete any unfinished contact investigation activities. Thus, this theme mapped to the *self-monitoring of behavior* BCT and *enablement* Behavior Change Wheel function.

Next, respondents described how feedback reports enabled them to learn about their own individual performance as well as aggregate performance of all CHWs at the clinic. One CHW explained:

> *“For me the [feedback] dashboards were fine and they used to remind us, for example, when you forget and you did not do the clinical evaluation…how are you performing and how was the clinic also performing*.*”*

Thus, through feedback reports, the CoP facilitated the *feedback on behavior* BCT and functioned through *enablement*.

CHWs suggested that feedback reports helped them gauge progress toward their objective of providing complete evaluation and linkage to treatment for all TB contacts. For example, one respondent explained:

> *“[Feedback reports] were good because they used to tell us what we should do, where we are delaying, what we are not doing well. Then we used to improve. They used to be good and in fact we should need them*.*”*

By drawing attention to discrepancies between their activities and goals, CHWs emphasized that feedback reports helped them “get back on track.” This theme mapped to the *discrepancy between current behavior and goal* BCT and the *enablement* Behavior Change Wheel function.

### Activity 2: Collaborative improvement meetings

CHWs described their experiences participating in weekly collaborative improvement meetings where they shared their experiences performing contact investigation, reviewed feedback reports with peers, and solved problems together. One CHW explained:

> *“I had worked before [the CoP was established] and they [the clinic staff] used to not know the challenges which we had and thus I used to spend three months on a problem. There is no way you could get over it. When I got the meetings weekly, I would share my ideas and problems and thus get a solution at that time*.*”*

This theme mapped to the *problem solving* BCT and *enablement* intervention function.

CHWs described that the feedback report discussions enabled them to support each other to reach their goal of completing contact investigation. For example, one respondent explained:

> *“Whenever you would get a challenge you could discuss it with other [CoP] members and they would give you advice. Because everyone gets their experience in a different way. We came to know that if this patient is not comfortable with me, I can switch to another community health worker. And things are sorted. So it was really good*.*”*

The CHWs recognized that each CoP member had particular experiences and skills that they could use to support each other. Thus, this theme mapped to the *social support (practical)* BCT and to the *enablement* intervention function.

CHWs also described that the collaborative improvement meetings enabled them to compare their own metrics against each other. The field notes indicated that the CoP discussed each member’s performance one-by-one. For example, weekly performance reports itemized and flagged incomplete contact investigation procedures as “missing forms.” One respondent explained:

> *“I would first rush and look at the pending [reports] I have, the missing forms I have. Then I look through and see my number and say “Ahh, I have two missing forms” and others have ten, others have four. Then you would say “Why do you have four, why do you have ten?” Then they would remind us of the missing people [household contacts] …Then when you go back, you call that home, you ask them what you were missing*.*”*

Because CHWs directly compared their own performance to that of their peers during group audit-and-feedback, this theme mapped to the *social comparison* BCT and *modelling* intervention function.

Furthermore, CHWs described that the collaborative improvement meetings increased their professional autonomy. The field notes described instances when CHWs brainstormed solutions for problems they were facing and then presented these solutions to the research coordinator to enact change. In interviews, CHWs explained that the CoP enhanced their decision-making power to propose and carry out innovations:

> *“They called us a team. Then they introduced to us what we were going to do and learnt that it was going to majorly depend on our side, as the [CoP] team. Because assuming we got a problem and needed a solution, we had to sit together and see the way forward. So, majorly it was on our decision making*… *In our teaching, we usually get orders from above. You are told what to do. And it’s not from down to up. But this time it was from down to up. So it wasn’t expected that way. It was new*.*”*

Many CHWs reported that having greater agency was motivating and enabled them to make meaningful changes to facilitate their work. This theme mapped to the *restructuring the social environment* BCT and *environmental restructuring* intervention function.

### Activity 3: Real-time communication among members

During weekly collaborative improvement meetings, CHWs concluded that having a system to communicate in real-time could facilitate timely support when problems arose in the field. The research team provided funding for and set up a WhatsApp messaging group to enable real-time communication between CoP members. The CHWs explained that having a WhatsApp group message gave them immediate access to a network of people should they experience a problem, especially in the field:

> *“If you needed some support, someone is there available for you to really support you with something that is challenging at the moment, which wasn’t there before*…*We developed a WhatsApp group. We used to communicate via phone calls and if one of the supervisors is not picking, [then] another is available. It really made some changes in the [communication] process*.*”*

This theme mapped to the *social support (practical)* BCT and *enablement* function.

### Activity 4: Didactic education sessions

CoP members invited experienced clinicians to deliver didactic education sessions on topics of interest. For example, field notes suggested that the CoP enabled members to identify gaps in their knowledge and skills for screening TB in children. CHWs then requested a didactic session on TB in children:

> *“I didn’t know how to screen TB in children. But during the CoP, we got the skills through our doctor*… *He gave us other skills of screening TB in children… It has raised me from one level of just being a community health worker of general health care to a more skilled [one] in TB…I can screen a child for TB, and I can give a full session on TB in children and adults*.*”*

The CHWs suggested that receiving educational sessions enabled them to improve their self-efficacy. This theme mapped to the *instruction on how to perform a behavior* BCT and *education* and *training* intervention functions.

Furthermore, CHWs shared that expanding their interpersonal and technical skills through the didactic sessions enabled them to work independently in the community as health workers, or *musawo*, similar to nurses and doctors. One respondent shared:

> *“[The CoP] taught me how to be patient with patients, it taught me how to be a humble person to patients and a loving person… sometimes it is not easy for a doctor or a nurse to go to a patient’s house but a CHW goes deep inside. And on the side of the patients, it makes them happy to see a musawo coming to his house sitting on that dirty chair, sitting on that dirty mat, it makes the patient happy*.*”*

CHWs emphasized that the trainings enabled them to identify themselves as more skilled and compassionate health workers, mapping to the *identity associated with changed behavior* BCT and the *training* and *education* intervention functions.

### Activity 5: Clinic-wide staff meetings

During collaborative improvement meetings, CoP members discussed how to improve communication between CHWs and clinic staff to facilitate their work. Thus, the CoP began holding meetings between CHWs and Kampala Capital City Authority (KCCA) clinic staff, including the clinic in-charge, lab personnel, TB leader, and clinicians. These meetings provided an opportunity for CHWs to discuss their contributions with clinic staff. One CHW described:

> *“We were very much recognized by the KCCA people and I think they even appreciated the work that was done*…*Before they used to not recognize community health workers very much. They could minimize [our work] a bit. But by that time at least some change was there. They recognized what was done in the community because we used to even refer some other people for other problems, not only TB and HIV*.*”*

Through clinic-wide staff meetings, clinic staff began recognizing CHWs’ work, improving their social standing within the clinic. This theme mapped to the *restructuring the social environment* BCT and *environmental restructuring* intervention function.

The CHWs explained that showcasing their successes to clinic staff also motivated clinic staff to support them when they faced barriers at the clinic. One respondent explained:

> *“They were all supportive, from sister in-charge to everyone, they were all supportive… in case I wanted anything, maybe from sister in-charge or lab or from a doctor or from a nurse, I could get it immediately…Because of the work I was doing. They saw that the work was good, they would make it easy for you*.*”*

Thus, this theme mapped to the *social support (practical)* BCT and *enablement* intervention function.

These meetings also created opportunities for clinic staff and CHWs to problem solve together to improve TB care in the clinic and community. One CHW described:

> *“Another motivation was that we were able to communicate in our meetings… We had a chance to sit with our medical team of our facilities… so that anything beyond our capability was able to be solved because we had the medical personnel with us as we were discussing or sharing our problems and challenges*.*”*

This theme mapped to the *problem solving* BCT and *enablement* function.

## Discussion

Previous studies of CoPs have suggested that they can improve performance by facilitating social support, knowledge sharing, knowledge creation, and formation of a professional identity (7, 9, 10). However, there remain critical gaps in characterizing the functional components and mechanisms of action of CoPs to improve delivery of health services in different settings, especially low-income countries (7, 9, 10). Our analysis contributes to the study of CoPs by characterizing the behavioral mechanisms described by members of a working CoP and locating them within a well-established taxonomy and model for characterizing behavior change interventions in implementation science. In so doing, we identified functional components of the CoP that elicited behavior change, which can be used for planning, implementing, and evaluating CoPs (22, 26).

We found that enabling CHWs to brainstorm and implement novel activities to improve their delivery of contact investigation was a core function of this CoP. Among the five key activities that CHWs perceived to facilitate their delivery of contact investigation, two were initially implemented by the research team and maintained by the CoP (i.e., review of feedback reports and collaborative improvement meetings), and three were proposed and maintained by the CHWs in response to their discussions in the collaborative improvement meetings (i.e., real-time communications among members, didactic education sessions, and clinic-wide staff meetings). By linking these activities to BCTs and intervention functions, we identified that the three CoP-initiated activities were important for improving CHW performance by facilitating social support in the field, restructuring the social environment within and outside the clinic, and receiving relevant education. Future studies that use CoPs as an implementation strategy should consider establishing a social environment that encourages members to develop and implement their own initiatives to facilitate their delivery of health services.

Although other studies of CoPs have not used this approach to explicitly link activities to BCTs or intervention functions, their findings do suggest that similar constructs may be important for CoP functioning. For example, a study of a CoP including physicians, nurses, and pharmacists to improve HIV care in Namibia found that knowledge of clinical HIV and self-efficacy to deliver HIV services increased, while professional isolation decreased, after implementation (27). These findings suggest that this CoP may have functioned through the *instruction on how to perform a behavior* and *social support* BCTs, similar to our findings. Another study of a CoP including CHWs and traditional healers focused on Buruli ulcer care in Cameroon found that the CoP enabled CHWs to have more autonomy in providing patient care, which led to improved social standing (28), consistent with the *restructuring the social environment* BCT. Similarly, CHWs in our study reported being motivated by the ways the CoP restructured the social environment and offered increased autonomy and efficiency in solving problems. Taken together, these findings suggest that health worker CoPs can facilitate delivery of high-quality care by enhancing members’ knowledge, skills, social support, and social status, which may be important motivators.

Our study provides preliminary insights into how CoPs are distinct from other group learning interventions, such as quality improvement collaboratives. Like CoPs, quality improvement collaboratives are multidisciplinary groups that meet to share experiences and identify barriers to performance to address them in real-time. However, quality improvement collaboratives employ a quality improvement or learning collaborative expert to lead the group and define its purpose, goals, and expectations (29-31). Alternatively, CoPs are intended to be run by the internal members and function through a shared sense of ownership (32). Our analysis found that having a sense of ownership was critical for the CoP to share and create knowledge and that this sense of ownership shaped CHWs’ social and professional identities. Thus, CoPs may realize enhanced and more sustained engagement from team members, compared with other group learning programs that rely on experts to lead the team. Furthermore, CHWs described that having autonomy to propose their own innovations was critical to improving TB care with low-cost strategies. The flexibility offered by the CoP enabled CHWs to implement these innovations in real-time. In these ways, CoPs are particularly well-suited for resource-constrained settings, where flexible, low-cost quality improvement strategies are needed. Conversely, quality improvement collaboratives are less flexible and can be costly, limiting their feasibility for facilitating quality improvement in low- and middle-income countries (31, 33-36). Lastly, CoPs can have evolving goals and definitions of success, which might promote enhanced buy-in, effectiveness, and sustainability compared with other collaborative improvement strategies. Additional studies should further explore distinctions between types of collaborative learning interventions and ultimately their impact on health service delivery.

Our study has some notable limitations. Because the interviews were conducted by research staff, social desirability bias might have influenced CHWs to describe activities in an overly favorable way. Given the nature of qualitative research, our study may not be generalizable to CoPs with different members, goals, and contexts. Furthermore, our study included a single CoP with eight members; thus, additional studies including multiple CoPs are warranted to validate, modify, and/or refute our findings. However, our study was strengthened by its use of an open-ended interview guide that allowed themes to emerge inductively from the voices of participants, strengthening the validity of our findings. By situating these themes within a widely used implementation science taxonomy and model, we identified functional components of CoPs that may be transferrable to other similar settings. This can support replication for evaluating the implementation and effectiveness of CoPs for contact investigation, and later potentially for adaptation and scale-up. Lastly, by triangulating our interview findings with field notes collected through observations, we aimed to mitigate social desirability bias and gain a comprehensive understanding of CoP activities.

## Conclusions

In summary, we identified BCTs and intervention functions through which a CoP facilitated the delivery of high-quality TB care by CHWs in Uganda. Additional empirical studies are warranted to validate and/or modify these proposed functional components to better understand how and under what conditions CoPs can be implemented to facilitate CHW-delivered health services. By using implementation science theory to expand the study of CoPs, we hope that future CoPs can be appropriately adapted to maximize effectiveness and sustainability.

## Data Availability

Data will be available upon request

## List of Abbreviations

BCT: Behavior Change Technique
CHW: Community Health Worker
CoP: Community of Practice
COREQ: Consolidated Criteria for Reporting Qualitative Research
NTLP: National TB and Leprosy Program
TB: Tuberculosis
TIDieR: Template for Intervention Description and Replication

## Declarations

### Ethics approval and consent to participate

Each participant provided verbal consent. The study was approved by the Makerere University School of Public Health Higher Degrees Research Ethics Committee and Yale University Human Investigation Committee approved the study.

### Consent for publication

Not applicable

### Availability of data and materials

The datasets used and/or analysed during the current study are available from the corresponding author on reasonable request.

### Authors’ contributions

JLD, MAH, and RH conceived the design of this study. JG and MAH created the interview guide and JG, EO, and PKT revised the interview guide. JG obtained verbal consent and conducted the interviews. RH conducted data analysis. All authors substantively contributed to the interpretation of the data based on the local context and the current implementation science literature. RH drafted the initial manuscript and all authors substantively revised the manuscript. All authors read and approved the final manuscript.

## Acknowledgements

We would like to acknowledge the participation of all the community health workers who participated in our study and provided their insights.

## References

1. Countdown to 2030 Collaboration. Countdown to 2030: tracking progress towards universal coverage for reproductive, maternal, newborn, and child health. Lancet. 2018;391(10129):1538–48.

2. Kruk ME, Gage AD, Arsenault C, Jordan K, Leslie HH, Roder-DeWan S, et al. High-quality health systems in the Sustainable Development Goals era: time for a revolution. Lancet Glob Health. 2018;6(11):e1196–e252.

3. Rowe AK, Rowe SY, Peters DH, Holloway KA, Chalker J, Ross-Degnan D. Effectiveness of strategies to improve health-care provider practices in low-income and middle-income countries: a systematic review. Lancet Glob Health. 2018;6(11):e1163–e75.

4. Kok MC, Dieleman M, Taegtmeyer M, Broerse JEW, Kane SS, Ormel H, et al. Which intervention design factors influence performance of community health workers in low- and middle-income countries? A systematic review. Health Policy Plan. 2015;30(9):1207–27.

5. Glenton C, Colvin CJ, Carlsen B, Swartz A, Lewin S, Noyes J, et al. Barriers and facilitators to the implementation of lay health worker programmes to improve access to maternal and child health: qualitative evidence synthesis. Cochrane Database Syst Rev. 2013(10):CD010414.

6. Lave J, Wenger E. Legitimate Peripheral Participation in Communities of Practice. In Situated Learning: Legitimate Peripheral Participation. Cambridge: Cambridge University Press; 1991.

7. Ranmuthugala G, Plumb JJ, Cunningham FC, Georgiou A, Westbrook JI, Braithwaite J. How and why are communities of practice established in the healthcare sector? A systematic review of the literature. BMC Health Services Research. 2011;11(273).

8. Wenger E. Communities of Practice and Social Learning Systems: the Career of a Concept. In: Blackmore C. (eds) Social Learning Systems and Communities of Practice. Springer, London 2010.

9. Li LC, Grimshaw JM, Nielsen C, Judd M, Coyte PC, Graham ID. Use of communities of practice in business and health care sectors: A systematic review. Implementation Science. 2009;4:27.

10. Ranmuthugala G, Cunningham FC, Plumb JJ, Long J, Georgiou A, Westbrook JI, et al. A realist evaluation of the role of communities of practice in changing healthcare practice. Implementation Science. 2011;6.

11. Kislov R, Harvey G, Walshe K. Collaborations for leadership in applied health research and care: lessons from the theory of communities of practice. Implement Sci. 2011;6:64.

12. Lewis CC, Boyd MR, Walsh-Bailey C, Lyon AR, Beidas R, Mittman B, et al. A systematic review of empirical studies examining mechanisms of implementation in health. Implementation Science. 2020;15:1–25.

13. Lewis CC, Klasnja P, Powell BJ, Lyon AR, Tuzzio L, Jones S, et al. From Classification to Causality: Advancing Understanding of Mechanisms of Change in Implementation Science. Front Public Health. 2018;6:136.

14. Kislov R, Pope C, Martin GP, Wilson PM. Harnessing the power of theorising in implementation science. Implementation Science. 2019;14.

15. Michie S, Van Stralen MM, West R. The behaviour change wheel: a new method for characterising and designing behaviour change interventions. Implementation science. 2011;6(1):1–12.

16. Michie S, Richardson M, Johnston M, Abraham C, Francis J, Hardeman W, et al. The behavior change technique taxonomy (v1) of 93 hierarchically clustered techniques: building an international consensus for the reporting of behavior change interventions. Ann Behav Med. 2013;46(1):81–95.

17. Centers for Disease Control and Prevention. Uganda Country Profile 2017 [November 18, 2020]. Available from: https://www.cdc.gov/globalhivtb/where-we-work/uganda/uganda.html.

18. Armstrong-Hough M, Turimumahoro P, Meyer AJ, Ochom E, Babirye D, Ayakaka I, et al. Drop-out from the tuberculosis contact investigation cascade in a routine public health setting in urban Uganda: A prospective, multi-center study. PLoS One. 2017;12(11).

19. Tong A, Sainsbury P, Craig J. Consolidated criteria for reporting qualitative research (COREQ): a 32-item checklist for interviews and focus groups. Int J Qual Health Care. 2007;19(6):349–57.

20. Timmermans S, Tavory I. Theory Construction in Qualitative Research: From Grounded Theory to Abductive Analysis. Sociological Theory. 2012;30(3):167–86.

21. Steinmo S, Fuller C, Stone SP, Michie S. Characterising an implementation intervention in terms of behaviour change techniques and theory: the ‘Sepsis Six’ clinical care bundle. Implementation Science. 2015;10(111).

22. Hoffmann TC, Glasziou PP, Boutron I, Milne R, Perera R, Moher D, et al. Better reporting of interventions: template for intervention description and replication (TIDieR) checklist and guide. BMJ. 2014;348:g1687.

23. Michie S, Abraham C, Eccles MP, Francis JJ, Hardeman W, Johnston M. Strengthening evaluation and implementation by specifying components of behaviour change interventions: a study protocol. Implementation Science. 2011;6:10.

24. Michie S, Atkins L, West R. The behaviour change wheel: a guide to designing interventions. London: Silverback Publishing; 2014.

25. Morse JM. The Significance of Saturation. Thousand Oaks, CA: Sage Publications; 1995.

26. Proctor EK, Powell BJ, McMillen JC. Implementation strategies: recommendations for specifying and reporting. Implement Sci. 2013;8:139.

27. Bikinesi L, O’Bryan G, Roscoe C, Mekonen T, Shoopala N, Mengistu AT, et al. Implementation and evaluation of a Project ECHO telementoring program for the Namibian HIV workforce. Hum Resour Health. 2020;18(1):61.

28. Awah PK, Boock AU, Mou F, Koin JT, Anye EM, Noumen D, et al. Developing a Buruli ulcer community of practice in Bankim, Cameroon: A model for Buruli ulcer outreach in Africa. PLoS Negl Trop Dis. 2018;12(3):e0006238.

29. Nadeem E, Weiss D, Olin SS, Hoagwood KE, Horwitz SM. Using a Theory-Guided Learning Collaborative Model to Improve Implementation of EBPs in a State Children’s Mental Health System: A Pilot Study. Adm Policy Ment Health. 2016;43(6):978–90.

30. Carter P, Ozieranski P, McNicol S, Power M, Dixon-Woods M. How collaborative are quality improvement collaboratives: a qualitative study in stroke care. Implement Sci. 2014;9(1):32.

31. Nadeem E, Olin SS, Hill LC, Hoagwood KE, Horwitz SM. Understanding the components of quality improvement collaboratives: a systematic literature review. Milbank Q. 2013;91(2):354–94.

32. Alcalde-Rabanal JE, Becerril-Montekio VM, Langlois EV. Evaluation of Communities of Practice performance developing implementation research to enhance maternal health decision-making in Mexico and Nicaragua. Implement Sci. 2018;13(1):41.

33. Nadeem E, Olin SS, Hill LC, Hoagwood KE, Horwitz SM. A literature review of learning collaboratives in mental health care: used but untested. Psychiatr Serv. 2014;65(9):1088–99.

34. Freney E, Johnson D, Knox I. Promoting Breastfeeding-Friendly Hospital Practices: A Washington State Learning Collaborative Case Study. J Hum Lact. 2016;32(2):355–60.

35. Zamboni K, Baker U, Tyagi M, Schellenberg J, Hill Z, Hanson C. How and under what circumstances do quality improvement collaboratives lead to better outcomes? A systematic review. Implement Sci. 2020;15(1):27.

36. Garcia-Elorrio E, Rowe SY, Teijeiro ME, Ciapponi A, Rowe AK. The effectiveness of the quality improvement collaborative strategy in low- and middle-income countries: A systematic review and meta-analysis. PLOS ONE. 2019;14(10):e0221919.

