## Supplemental Files for "Functional components of a community of practice to improve community health worker performance: A qualitative study"

**Additional File 1. Interview guide**

*Domain 1: Perception of role*

1. Looking at your current job working as a community health worker following a Community of Practice approach on the pilot study, is the job what you expected?

*Probes*:

How does it make you feel?

Does it make you excited?

2. In our previous discussion, we talked about what it means to be a community health worker, has that changed over time?

*Probes*:

What is it really like being a community health worker?

3. What has kept you encouraged (motivated) to continue doing community health worker activities?

*Probes*:

What parts of your work give you motivation?

4. In the last six months, has being a community health worker changed you in any way?

*Probes*:

How so?

How has being a CHW affected your life?

5. Which aspects of your job do you consider to be the most important? (e.g. collecting good quality sputum, delivering Xpert results to clients, doing HIV tests, etc.)

*Domain 2: Experiences during the Community of Practice pilot study*

6. What have been your experiences during the pilot study working with other departments at your clinic?
7. Has your experience working with the pilot changed your relationship with others at the clinic?
8. How did you find support for your work from others at your clinic?
9. Can you tell me about a time one of your fellow CHWs helped you?

*Probes*:

How did they help?

How did you ask them for help?

10. Do you look up to any of your colleagues? If yes, who and why?

*Domain 3: Feedback reports*

11. How do you find the dashboards?

*Probe:*

Which part(s) of the report do you find most useful?

12. When you are given a weekly report, what do you look at first?

*Domain 4: Thank you and conclusion*

13. Is there anything else you want to tell me about being a CHW?
14. Is there any question I didn’t ask that I should have asked?

**Additional File 2. Contextual factors that facilitated Community of Practice activities**

| **Community of Practice Activity** | **Contextual Factor(s)** | **Quotation** |
| --- | --- | --- |
| Individual review of feedback reports | The tablet reader and electronic mode of delivering the feedback reports, once adopted, provided a more timely and readily accessible way for Community of Practice members to receive feedback. | “Because we were able to get our reports without moving from our work places, because whatever we were doing was directly reported, the paper [report] was not so much needed. We were able to give in our reports as soon as we have finished our client, a report was directly sent to the server so there was an easy follow [up] assuming there was something which did not go well. And at least there was easy communication to correct the problem or mistake instead of waiting or calling, you were able to get out of that dilemma as soon as it happened.” |
| Collaborative improvement meetings | The electronic feedback reports on the tablets enabled Community of Practice members to easily review each other's reports for discussion during the meetings. | “So you could be talking [in the CoP meetings] and the then the person there could follow whatever was being discussed and then scroll in time... You know if you are sharing a soft [electronic] copy it is easier. They could share it with these people through email so anyone could have it soon and they would love it.” |
| Real-time communications among members | Access to a mobile data plan and supportive supervisors facilitated communication among Community of Practice members and supervisors in the field. | “First of all, our supervisors used to help us in all areas where we could get challenges. They were always readily available. You could call them whether you were in a home visit and they could say no, you can do this or that, so they were always there for us. They could also provide us with timely transport to go to those home visits. We were very well facilitated. So it could make work easy for me to plan with a patient and visit as soon as possible.” |
| Didactic education sessions | None | N/A |
| Clinic-wide staff meetings | Supportive supervisors advocated for Community of Practice members at the clinic. | “You would call [CoP supervisors] if something is not done [at the clinic] and they would come to Kawaala [Health Center] and we would have a meeting on the issues.” |
